# Nasopharyngeal and serological anti SARS-CoV-2 IgG/IgA responses in COVID-19 patients

**DOI:** 10.1101/2021.01.13.20249038

**Authors:** Bernadette Crescenzo-Chaigne, Sylvie Behillil, Vincent Enouf, Nicolas Escriou, Stephane Petres, Marie Noelle Ungeheuer, Jade Ghosn, Sarah Tubiana, Lila Bouadma, Sylvie van der Werf, Caroline Demeret, for the French COVID cohort study group

## Abstract

**Background:** The systemic antibody responses to SARS-CoV-2 in COVID-19 patients has been extensively studied. However, much less is known about the mucosal responses in the upper airways at the site of initial SARS-CoV-2 replication. Local antibody responses in the nasopharyngeal epithelium, that are likely to determine the course of infection, have not been analysed so far nor their correlation with antibody responses in serum.

**Methods:** The IgG and IgA antibody responses were analysed in the plasma as well as in nasopharyngeal swabs (NPS) from the first four COVID-19 patients confirmed by RT-qPCR in France. Two were pauci-symptomatic while two developed severe disease. Taking advantage of a comprehensive series of plasma and nasopharyngeal samples, we characterized their antibody profiles from the second week post symptoms onset, by using an in-house ELISA to detect anti-SARS-CoV-2 Nucleoprotein (N) IgG and IgA.

**Results:** Anti-N IgG and IgA antibodies were detected in the NPS of severe patients. Overall, the levels of IgA and IgG antibodies in plasma and NPS appeared specific to each patient.

**Conclusions:** Anti-N IgG and IgA antibodies are detected in NPS, and their levels are related to antibody levels in plasma. The two patients with severe disease exhibited different antibody profiles that may reflect different disease outcome. For the pauci-symptomatic patients, one showed a low anti-N IgG and IgA response in the plasma only, while the other one did not exhibit overt serological response.

## Introduction

The Coronavirus disease 2019 (COVID-19) is a respiratory disease that has been threatening human health since early 2020, causing a global pandemic with dramatic social and economic consequences. COVID-19 is caused by severe acute respiratory syndrome coronavirus 2 (SARS-CoV-2), a highly transmissible and pathogenic coronavirus that emerged in late 2019 and has spread rapidly around the world during the year 2020 [1]. Considerable efforts are being made to understand SARS-CoV-2 epidemiology and pathogenesis, which is crucial for antiviral drug development and vaccine design [2,3].

Detecting SARS-CoV-2 RNAs by RT-qPCR from nasopharyngeal swabs (NPS) is a standard approach for COVID-19 diagnosis. A detailed characterization of the immune response to SARS-CoV-2 infection is mandatory, to understand its contribution to virus clearance and establishment of protection.

The first point of SARS-CoV-2 entry are the mucosal surfaces of the upper respiratory tract [4]. Accordingly, characterization of the immune response in mucosal samples of infected patients, compared to the humoral response in plasma, is of interest. IgA have important roles in the immune response of mucosal surfaces [5]. Indeed, the antibody pool in the mucosa contains a higher proportion of IgA than in the plasma, and IgA may be important contributors to protection against viruses that target the mucosal surfaces [6]. IgA antibodies directed against SARS-CoV-2 are produced rapidly after infection and remain elevated in the serum for about one month after the onset of symptoms [7–10]. The levels of IgA proved significantly higher in patients with severe compared to mild or moderate infection, following the same trend as anti SARS-CoV-2 IgG [7,9]. Antibodies binding to the Receptor Binding Domain (RBD) of the Spike protein can neutralize SARS-CoV-2. Significantly, anti RBD IgA have been shown to have more potent neutralizing activities than IgG [11,12]. However, the contribution of IgA in the mucosal antibody-mediated response to SARS-CoV-2 has not been reported. The mucosal response to SARS-CoV-2 has been assessed in the saliva of COVID-19 patients, while mucosal immunity in the NPS has not been characterized although it is the mucosal surface of the initial site of viral replication. Comparing anti-N IgA and IgG antibody levels of patient’s NPS to their levels in plasma provides a more complete picture of the antibody-mediated immune response to SARS-CoV-2 infection.

In this study, we used an in-house ELISA [13] to detect IgG and IgA directed against the SARS-CoV-2 Nucleoprotein (N) in the NPS and in the plasma of four of the first recognized COVID-19 patients in France [14]. The patients experienced different clinical evolutions, two exhibiting severe disease and two mild and pauci-symptomatic disease. This set of patients is particularly valuable owing to (i) the different disease progression between patients; (ii) the extensive sampling over the COVID-19 illness period. It provides a unique opportunity to compare the systemic and mucosal antibody response in severe patients during the course of illness and in pauci-symptomatic individuals until and after their full recovery.

We detected IgG and IgA antibodies in the NPS of the two severe patients, demonstrating a mucosal antibody response to SARS-CoV-2. Serological and mucosal antibody profiles were specific to each patient, with higher antibody levels in the plasma of the severe patients as previously reported [15][16] that were also reflected in the nasopharyngeal mucosa. Our study highlights a diversified humoral response to SARS-CoV-2 infection, with IgA to IgG ratios both in blood and NPS potentially underlying different disease outcome.

## METHODS

### Study design

The objective of this study was to analyse the levels of anti-N SARS-CoV-2 IgA and IgG antibodies in the NPS and in the plasma of four confirmed COVID-19 cases with different clinical histories. We used an anti-N ELISA, the performance of which was evaluated using NPS and serum from pre-epidemic individuals.

### Cohorts

Pre-epidemic NPS originated from the National Reference Center (NRC) for respiratory viruses and were confirmed to be SARS-CoV-2 negative by RT-PCR (data not shown). Pre-epidemic sera were provided by the ICAReB platform (Clinical Investigation and Access to Research Bioresources, Institut Pasteur) or the Center for translational Science, Institut Pasteur. All pre-epidemic samples were collected before November 2019. Samples from donors were collected in accordance with local ethical guidelines by Pasteur Institute.

The NPS and plasma samples for the four COVID-19 patients are part of patients included in one of the participating center (i.e., Hôpital Bichat, APHP, Paris, France) of the French COVID cohort assessing hospitalized patients with a virologically-confirmed COVID-19 (NCT04262921) [17]. This study was conducted with the understanding and the consent of each participant or its surrogate. Ethics approval was obtained from the French Ethic Committee CPP-Ile-de-France VI (ID-RCB: 2020-A00256-33). NPS and plasma or sera were heat-inactivated 30 min at 56°C

### ELISA-N

The ELISA test was performed as described in Grzelak et al [13]. Briefly, 96-well ELISA plates were coated overnight with 50 ng of purified bacterially expressed N protein of SARS-CoV-2 in PBS. After washing 3 times with PBS-0.1% Tween 20 (PBST), 200μl of PBST-3% milk were added and incubated 1 h at 37°C. After washing, 100 μl of diluted plasma or NPS in PBST-3% milk were added and incubated 1 h at 37°C. After washing, plates were incubated with 5,000 or 4,000-fold diluted peroxidase-conjugated goat anti human IgG or IgA respectively (#2040-05 and #2050-05 -Southern Biotech) for 1h. Plates were revealed by adding 100 μl of HRP chromogenic substrate (TMB, Eurobio Scientific) after 3 washing steps in PBST. After 10 min incubation, the reaction was stopped by adding 1M H_3_PO_4_ and optical densities were measured at 450 nm (OD 450). All tests were performed in triplicate.

## RESULTS

### Nasopharyngeal swabs and blood samples

Nasopharyngeal swabs and serum samples collected before November 2019 were used as negative samples (pre-COVID-19 pandemic). To investigate the kinetics of anti SARS-CoV-2 mucosal antibody responses in comparison with systemic responses, we took advantage of the comprehensive longitudinal sampling performed for four of the first COVID-19 cases detected in France.

The patients were admitted at Hospital Bichat, Paris, and confirmed for SARS-CoV-2 infection by RT-PCR at the time of admission. They experienced different clinical evolutions: two exhibited severe disease, one being fatal, and two had mild disease [14]. Three to eight NPS samples collected between days 10 and 24 post symptoms onset (PSO), and five to nine blood samples collected between days 10 and 32 PSO, including dates matching the NPS, were used to explore the IgG and IgA anti-N response of each of the patient. Clinical history and sampling periods are summarized in Table 1.

**Table 1.** Summary of sampling dates along with patient’s illness history. The data are from (14).

| | day PSO* | illness history** | SARS CoV2<br>RT-qPCR<sup>\$</sup> | NPS<sup>\$\$</sup> | plasma<sup>\$\$\$</sup> |
| --- | --- | --- | --- | --- | --- |
| patient 1 | 0 |  |  |  |  |
|  | 1 | influenza like symptom |  |  |  |
|  | 6 | hospital admission, diagnosed COVID-19 | PCR + |  |  |
|  | 10 | ICU admission, symptoms worsening | PCR - |  |  |
|  | 13 | ICU discharge, patient improvement | PCR - |  | X |
|  | 14 | PCR neg | PCR - | X |  |
|  | 19 | asymptomatic |  | X | X |
|  | 21 | asymptomatic |  | X | X |
|  | 23 | asymptomatic |  | X | X |
|  | 25 | hospital discharge |  |  | X |
|  | 32 |  |  |  | X |
| patient 3 | 0 |  |  |  |  |
|  | 1 | fever and diarrhoea |  |  |  |
|  | 4 | hospital admission- Not diagnosed as COVID19 |  |  |  |
|  | 5 | ICU admission- acute respiratory failure |  |  |  |
|  | 7 | ICU-diagnosed COVID19 | PCR + |  |  |
|  | 14 |  |  | x |  |
|  | 15 | ICU |  | x | x |
|  | 16 | ICU |  | x | x |
|  | 17 | ICU |  |  | x |
|  | 18 | ICU |  | x | x |
|  | 19 | ICU |  | x | x |
|  | 20 | ICU | PCR + | x | x |
|  | 21 | ICU |  | x | x |
|  | 22 | ICU | PCR - |  | x |
|  | 24 | death | PCR - | x | x |
| patient 4 | 0 |  |  |  |  |
|  | 1 | moderate influenza-like symptoms |  |  |  |
|  | 2 | hospital admission-diagnosed COVID19 | PCR + |  |  |
|  | 3 | mild symptoms | PCR + |  |  |
|  | 8 | mild symptoms | PCR + |  |  |
|  | 9 | mild symptoms | PCR - |  |  |
|  | 11 | asymptomatic |  |  |  |
|  | 12 | asymptomatic | PCR - |  |  |
|  | 14 | asymptomatic |  | x | x |
|  | 16 | asymptomatic |  | x | x |
|  | 18 | asymptomatic |  |  | x |
|  | 19 | asymptomatic |  | x |  |
|  | 20 | asymptomatic |  |  | x |
|  | 21 | hospital discharge |  |  |  |
|  | 28 | asymptomatic |  |  | x |
| patient 5 | 0 |  |  |  |  |
|  | 1 | hospital admission-mild symptoms- dry cough |  |  |  |
|  | 2 | PCR +-high viral load--COVID diagnosed | PCR + |  |  |
|  | 3 | mild symptoms | PCR + |  |  |
|  | 8 | asymptomatic | PCR + |  |  |
|  | 10 | asymptomatic | PCR - | x | x |
|  | 12 | asymptomatic |  | x | x |
|  | 14 | asymptomatic | PCR - | x | x |
|  | 16 | asymptomatic | PCR - |  | x |
|  | 21 | hospital discharge |  |  |  |
|  | 23 | asymptomatic |  |  | x |

\* days post symptoms onset, \*\* main steps in clinical evolution, \$ Detection of SARS CoV2 viral RNA RT-qPCR in NPS, \$\$ NPS samples used in this study, \$\$\$ plasma samples used in this study.

### Detection of anti-N antibodies by ELISA

We previously developed an anti-N ELISA test to assess antibody level in plasma or sera [13]. In this study, we adapted this ELISA to the detection of anti-N antibodies in the NPS, and used it to compare the production of antibodies in the plasma and in the NPS. Serial 2-fold dilutions of the samples were assessed for anti-N IgG and IgA antibodies, using anti IgG and anti IgA secondary antibodies in otherwise strictly identical experimental settings. Sera or NPS samples of pre-epidemic patients were run in parallel to patient samples, to give the background anti-N IgG and IgA detection level in each fluid type.

Signals of both IgG and IgA anti-N antibodies (OD450) in the patient’s plasma were clearly above that observed for pre-pandemic samples for all patients except patient 5 (Figure1).

**Figure 1.**
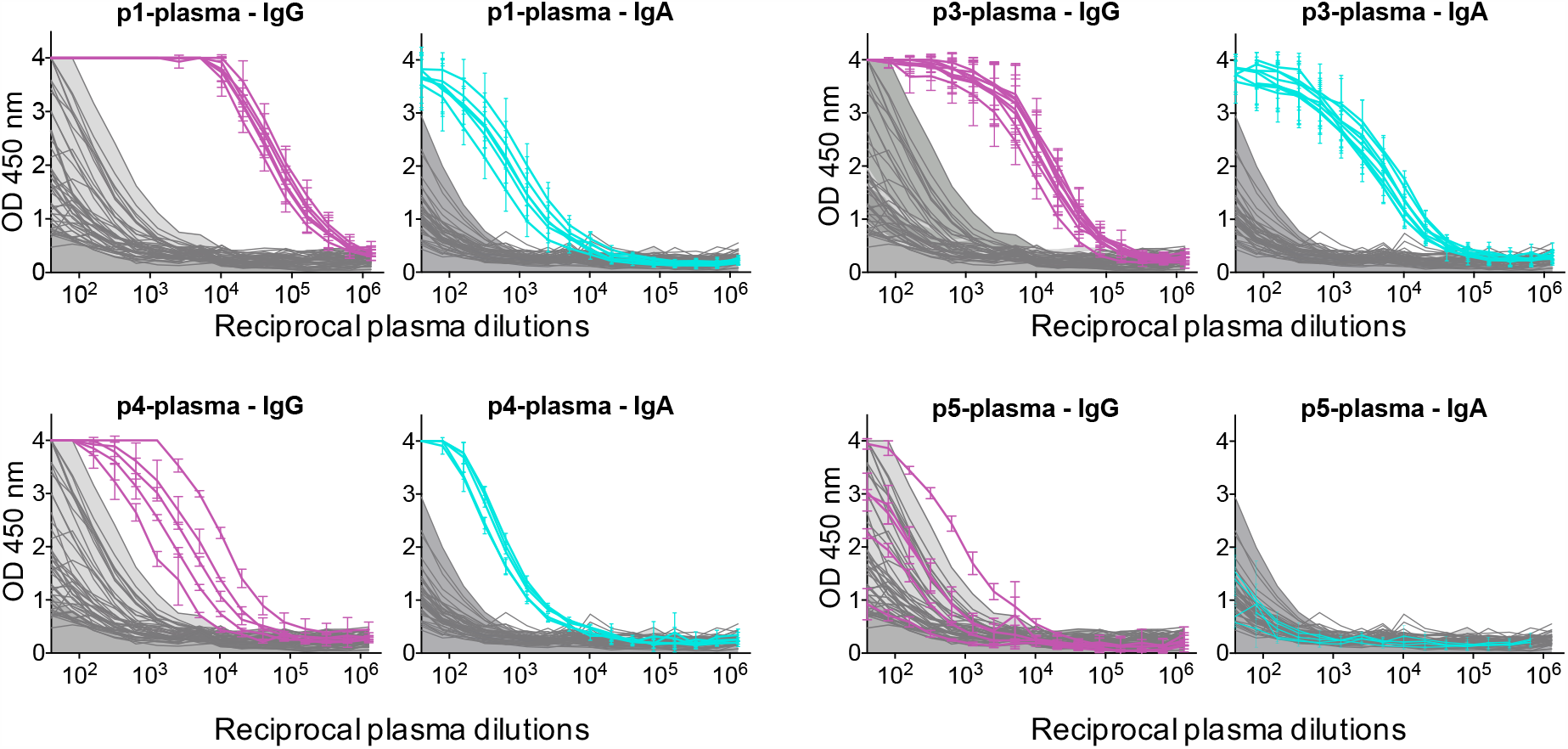
Detection of IgG and IgA anti N antibodies in patient’s plasma. ELISA measuring plasma IgG and IgA reactivity to SARS-CoV-2-N protein for each patient (p1,3,4,5). Graphs show the optical density units at 450 nm (Y axis) and reciprocal plasma dilutions (X axis) of each sample, taken at different times PSO, as described in Table 1. Dilutions of pre-epidemic control sera are in grey, the filled light grey area shows ELISA signals generated by negative serum samples.

Anti-N signals in the NPS were distinct from the pre-pandemic NPS for the two severe patients, indicating that anti-N IgG and IgA antibodies can be specifically detected in NPS samples from COVID-19 patients (Figure 2).

**Figure 2.**
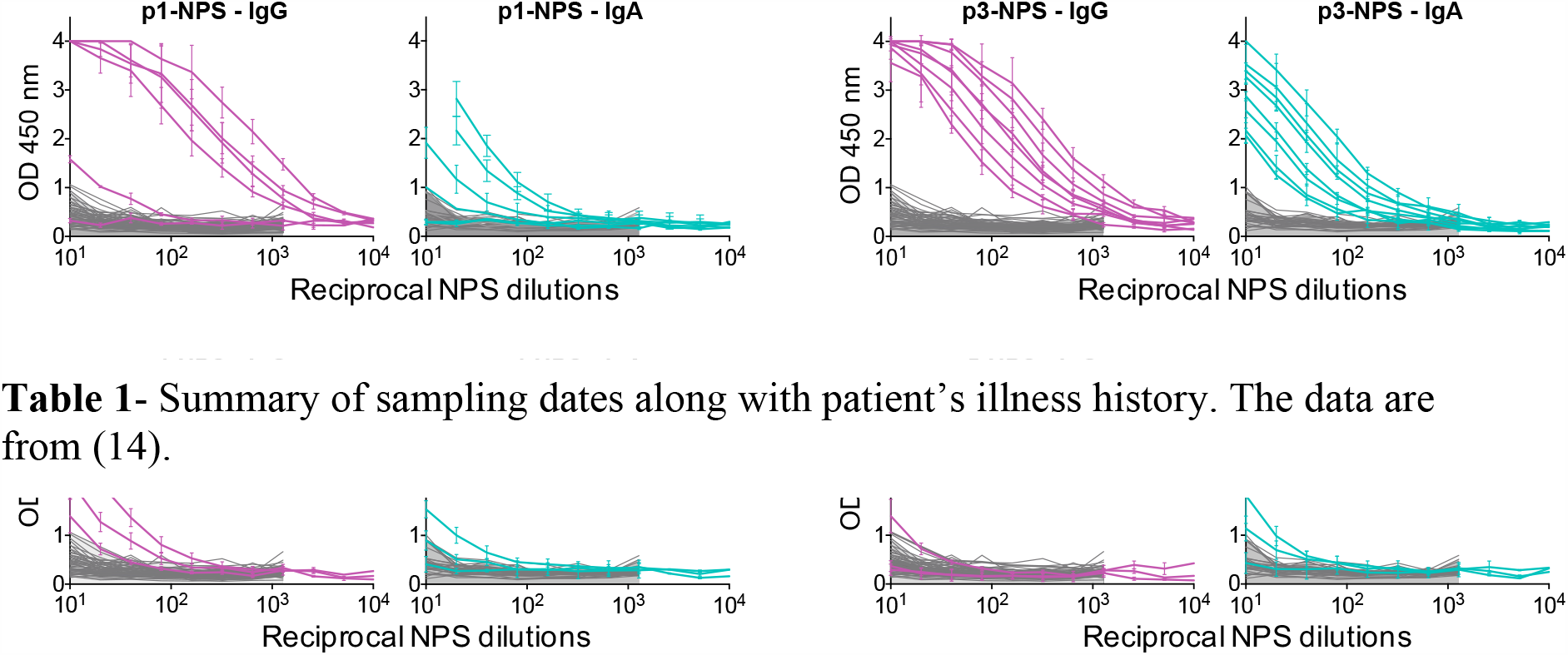
Detection of IgG and IgA anti N antibodies in patient’s NPS. ELISA measuring the IgG and IgA reactivity in the NPS for each patient (p1,3,4,5). Graphs show the optical density units at 450 nm (Y axis) and reciprocal plasma dilutions (X axis) of each sample, taken at different times PSO, as described in Table 1. Dilutions of pre-epidemic control NPS are in grey, the filled light grey area shows ELISA signals generated by negative NPS samples.

Antibody titers were derived from the dilution curves, and expressed as RD50 (*i.e* the reciprocal serum dilution necessary to obtain 50% of maximum ELISA OD450 values). For both IgG and IgA measurements, matched cut off values were calculated for the plasma and NPS by calculating the mean RD_50_ titer of negative pre-pandemic samples + 3 Standard deviations (sera: n=46; NPS: n=48 for IgG; n= 27 for IgA).

### The anti N serological response of the patients

**Patient 1**, who developed secondary severe symptoms, showed a robust serological response all along the sampling period, from the patient’s discharge from ICU (day 12 PSO) until day 32 PSO after the patient’s full recovery and discharge from hospital (Table 1). The IgG levels were constantly high (47.000 to 94.000 RD50, cut off value = 379), and the IgA levels were low (360 to 1300 RD50, cut off value = 56) (Figure 3). Patient 1 therefore demonstrated markedly different levels of IgG and IgA in the plasma, with IgG titers 70- to 130-fold higher than IgA titers. The humoral response in the plasma of this patient appears prominently driven by IgG, and persists over time without any obvious changes.

**Figure 3.**
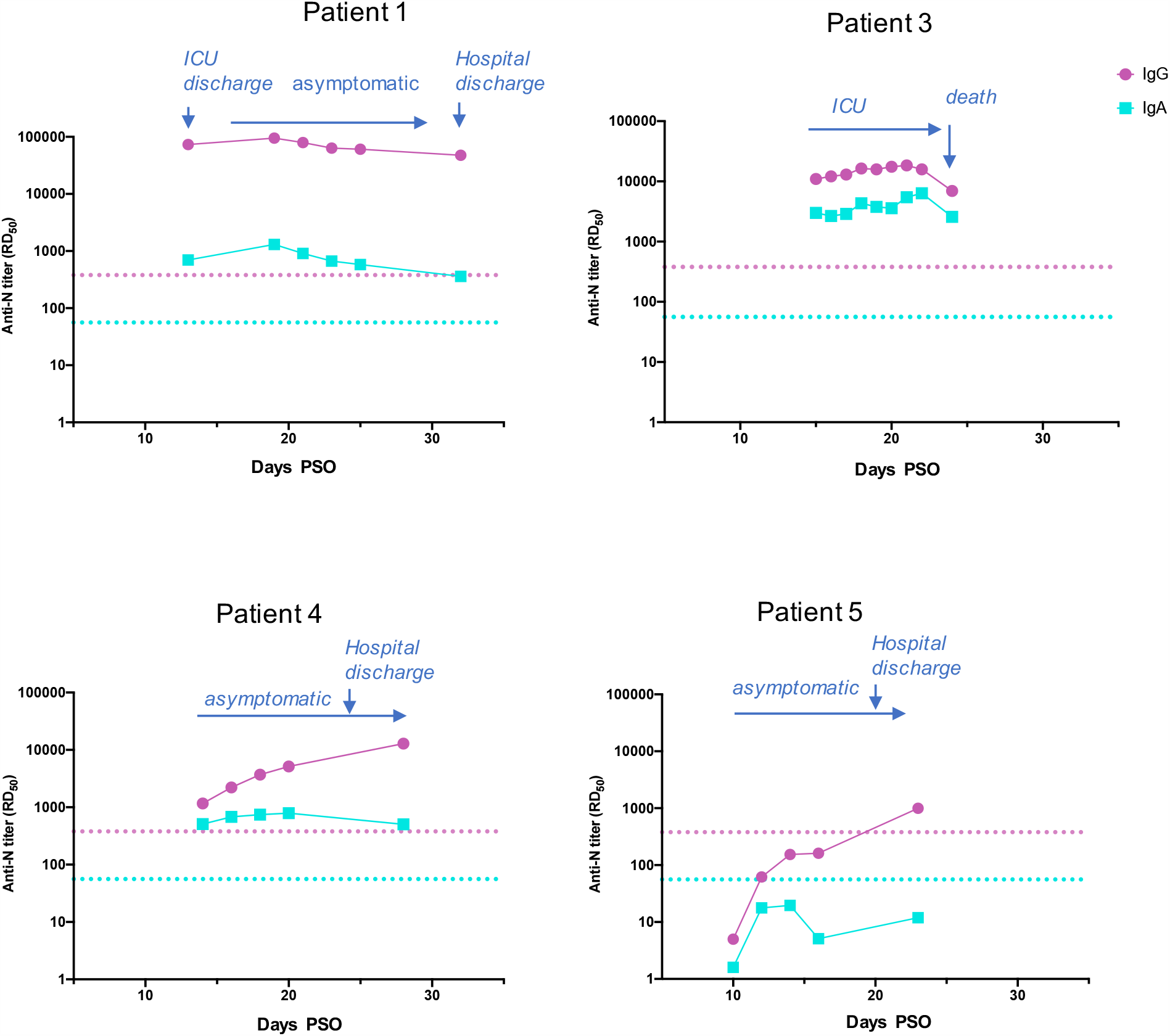
IgG ang IgA antibody titers in patient’s plasma. Antibody titers calculated as RD_50_ (Y axis) are plotted according to the day PSO (X axis). Main steps of the patient’s clinical history are shown by arrows. The positive thresholds of IgG and IgA detection, deduced from the mean IC_50_ of pre-epidemic samples + 3SD, are shown as pink (IgG) and blue (IgA) dotted lines respectively.

**Patient 3**, with a rapidly progressing severe disease leading to death, had lower IgG levels but higher IgA levels compared to Patient 1. Indeed, between days 15 and 22 PSO, IgG titers ranged from 11,000 to 15,000 RD50, and IgA titers from 2,600 to 6,400 RD50 (Figure 3), and IgG levels were only 2.5- to 4.5-fold higher than IgA titers. Antibody levels did not vary significantly until a decline on day 24 PSO, when patient 3 died, probably reflecting the general organism breakdown.

P**atient 4** had IgG antibody levels progressively increasing over time in the plasma (from 1,000 to 12,000 RD50). In contrast, the levels of IgA antibodies were constant and low, at titers around 600 RD50 (Figure 3). The IgG/IgA ratio increased from 2.5 to 6.5 from days 14 to 20 PSO, and reached a value of 25 by day 28 PSO. This reflects a developing serological IgG response after patient’s recovery.

**Patient 5** did not show any significant seroconversion to SARS-CoV-2 N protein, despite development of mild symptoms and confirmation of infection by RT-PCR [14]. For this patient IgG antibodies were only detected at a titer above the cut off value at day 23 PSO (Figure 3), after discharge from the hospital and full recovery.

### Detection of anti N antibodies in nasopharyngeal swab

Since, unlike plasma, the amount of biological material recovered in NPS and placed in liquid transport medium is not constant from one sample to another, we compared the IgG and IgA levels within each NPS sample, but did not perform day-by-day nor patient-to-patient comparisons. IgG and IgA ELISA signals above the cut-off were detected in NPS of **patients 1 and 3**, indicating the occurrence of a humoral response in the nasopharyngeal mucosa of these patients (Figure 4). The IgG to IgA ratio in NPS of **patient 1** markedly differed over time. It decreased from 80 at day 14 PSO to 9 and 14 at days 19 and 21 PSO, respectively. Then a sharp increase was observed at day 23, reflecting the collapse of IgA levels while IgG antibodies remained at high levels (Figure 4). Such IgA drop in the NPS occurred one week after the patient became free of symptoms (Table 1). **Patient 3** exhibited an IgG/IgA ratio in NPS oscillating between 4 and 8 over time. No evolution of this ratio was observed along with illness severity, which led to death on day 24. **Patient 4** showed a weak IgG response in the NPS at days 16 and 19 PSO, indicating a moderate but significant activation of the humoral response in the nasal mucosa, driven by IgG antibodies. **Patient 5** did not demonstrate any IgG response in NPS. As both patients 4 and 5 are devoid of anti N IgA in NPS, the IgG/IgA ratio is irrelevant.

**Figure 4.**
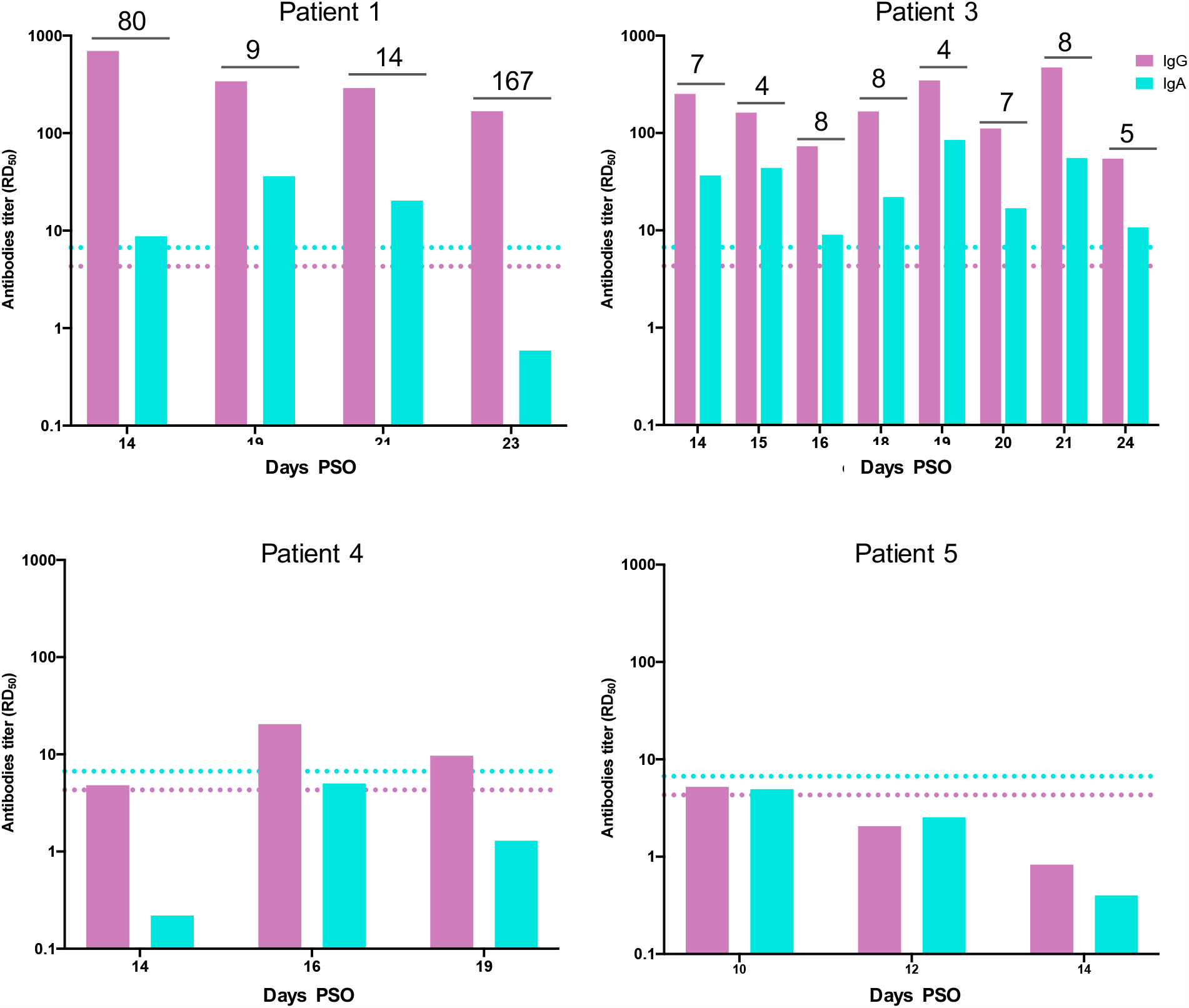
IgG ang IgA antibody titers in patient’s NPS. Antibody titers calculated as RD_50_ (Y axis) are plotted according to the day PSO (X axis). The positive thresholds of IgG and IgA detection, deduced from the mean IC_50_ of pre-epidemic NPS samples + 3SD, are shown as pink (IgG) and blue (IgA) dotted lines respectively. The ratio of IgG to IgA titers is indicated when appropriate.

## DISCUSSION

We report here the matched detection of SARS-CoV-2 anti-N IgG and IgA, in the NPS and in the plasma of four well-characterized COVID-19 patients, two severe patients, and two pauci-symptomatic patients who remained under survey at the hospital despite their mild symptoms given that the epidemic was in its early stage.

The two patients with severe disease, patients 1 and 3, showed stable anti-N IgG and IgA responses in the plasma, indicating that the maximal serological response had been reached, which is in line with the time post symptoms sampled [18]. Their serological response was nevertheless clearly distinct. The humoral response in the plasma of patient 1 was dominated by a strong IgG response, the IgG/IgA ratio being constantly around 100. Patient 3 had a less robust IgG response and a stronger IgA response than patient 1, with an IgG /IgA ratio between 2.5 and 4.5. Given that virus clearance and COVID-19 recovery occurred for patient 1 but not patient 3, who suffered an active viral replication until the fatal outcome of his disease, these results suggest that serological IgA antibodies are not the main drivers of disease resolution. We could also detect IgG and IgA in the NPS of these severe patients, indicating the occurrence of a humoral response at the initial site of infection, the nasopharyngeal mucosa. The IgA levels in NPS were consistently lower than IgG just as in plasma, suggesting that the mucosal response was dominated by an IgG response at late times of infection. This differs from the mucosal response that has been detected in COVID-19 patient’s saliva earlier post infection, where higher proportions of IgA were detected [19].

The mucosal IgA response of patient 1 developed after symptoms improvement following the severe phase of his illness. The IgG/IgA ratio in the NPS followed a v-shaped progression, dropping from 80 to about 10 due to a transient rise in the IgA levels, then back up to 100 owing to IgA collapse. The transient rise in mucosal IgA antibodies between 19 and 21 days PSO indicates the development of an IgA-based mucosal response, that is delayed compared to the serological response, since at the exact same days PSO the IgA levels in the plasma progressively decreased (Figures 3 and 4). Patient 3’s IgG/IgA ratio is constant over time, and in the order of 5 in both plasma and NPS. Beyond patient’s age, the antibody rate in the plasma, far higher for patient 1 than 3, could underlie different disease outcome. Patient 1 is characterized by a clear predominant IgG response in both plasma and NPS, detected upon disease resolution after the period of symptoms worsening. In contrast, patient 3’s IgG response remained moderate and close to the IgA levels both in the plasma and in the nasal mucosa, suggesting that the anti-N IgG humoral response might be one determinant of virus clearance. For the two pauci-symptomatic patients, while their clinical profiles were very similar (mild symptoms, full recovery at hospital discharge that corresponds to the last day or sampling), we also observed divergent anti-N humoral responses. Patient 4 exhibited an anti IgG response in the plasma still increasing at 28 days PSO, which reflects a delayed IgG response compared to severe patients, in agreement with previous reports [15]. Patient 4 seems to have developed an earlier serological IgA response, reflected by constant IgA levels in the plasma between days 16 and 20 PSO, although antibody measurement in earlier samples would be required to confirm that point. In addition, the serological IgA response of patient 4 appears transient, as the IgA level decreased on the last day of sampling (day 32 PSO), while the IgG level continued to increase. A sustained stable or increasing IgG response over time in symptomatic individuals with mild disease has recently been linked to a shorter time to disease resolution [20], as was the case of patient 3. We did not detect any IgA in the NPS of this patient, making the IgG/IgA ratio irrelevant. A weak and transient IgG response could be detected in the NPS of this patient during the asymptomatic period, concomitant with the increase of the IgG response in the plasma.

Patient 5 did not develop any significant antibody response against N, neither in the plasma nor in NPS. Such different humoral responses could not be due to a different sampling time, since they covered a similar period post symptoms for both pauci-symptomatic patients (days 14-28 and 10-23 for patients 4 and 5 respectively). The case of patient 5 further demonstrates that a RT-PCR confirmed COVID-19 infected individual may show a delayed seroconversion, as IgG were only detected at day 23 PSO. However, analysis of later time points would have been required to document a further increase in IgG titers, as shown for individuals with low initial antibody titers [21], or whether this patient belongs to the small fraction of pauci-symptomatic patients defined as “IgG non-responders” [15]. We extend here these observations to the IgA serological response, and show herein that patient 5 did not either develop any detectable mucosal humoral response. The fact that some pauci-symptomatic patients may not develop any humoral response neither in the nasal mucosa nor in serum, should be taken into account in seroprevalence surveys. It also suggests that non-humoral, innate or cell-mediated immunity may be sufficient to resolve infection as previously reported [22].

In conclusion, we compared for the first time the IgG and IgA responses in the plasma and in the NPS. Exploring four well-documented confirmed Covid-19 patients with different disease profiles, we identified different antibodies responses according to (i) the severity of the symptoms (symptomatic vs pauci-symptomatic) (ii) disease progression (severe leading to death vs severe resolved), (iii) variations among pauci-symptomatic patients. These conclusions are nevertheless to be substantiated with other longitudinal studies of both severe, pauci-symptomatic or asymptomatic individuals.

## Data Availability

The authors confirm that the data supporting the findings of this study are available within the article.

## Author contributions

BCC handled laboratory logistics and generated data. BCC and CD conceived the project, analysed and summarized the data, and drafted the manuscript. SB and VE shared samples and data for the analysis. JG, ST, LB provided the specimens and sera for the analysis. SP and NE provided the purified N, critical for the study. SW provided input on interpretation of the results and critically reviewed the manuscript. All authors reviewed and the article.

## ACKNOWLEDGMENTS

This work was supported by the ‘URGENCE COVID-19’ fundraising campaign of Institut Pasteur, by REACTING (Research & Action Emerging Infectious Diseases) and by the H2020 project 101003589 (RECOVER). The French Covid Cohort is funded through the Ministry of Health and Social Affairs (PHRC n°20-0424) and Ministry of Higher Education and Research dedicated COVID19 fund. Funding sources are not involved in the study design, data acquisition, data analysis, data interpretation or manuscript writing. List of the membership of the French COVID cohort study group : Mélanie RORIZ, Patrick RISPAL, Sarah REDL (CHG d’Agen), Laurent LEFEBVRE, Pascal GRANIER, Laurence MAULIN (CH du Pays d’Aix, Aix en Provence), Cédric JOSEPH, Julien MOYET, Sylvie LION-DAOLIO (CHU Amiens Nord), Rafael MAHIEU, Alexandra DU-CANCELLE, Vincent DUBEE (CHU d’Angers), Stéphane SALLA-BERRY, Aldric MANUEL, Gabriel MACHEDA, Mylène MAILLET, Bruno CHANZY (CH d’Annecy Genevois), Jean-Charles GAGNARD (Hôpital privé d’Antony), Guillermo GIORDANO, Clara MOUTON PERROT, Vincent PESTRE (CH Henri Duffaut, Avignon), Cécile FICKO, Marie GOMINET, Aurore BOUSQUET (Hôpital d’instruction des armées Begin), Charline VAUCHY, Kévin BOUILLER, Maïder PAGADOY, Solène MARTY-QUINTERNET, Quentin LEPILLER (CHU Jean Minjoz, Besançon), Cyril LE BRIS, Benoit THILL, Marie-Laure CASANOVA, Georges LE FALHER, Eric OZIOL (CH de Béziers), Hugues CORDEL, Nathalie DOURNON, Olivier BOUCHAUD, Ségo-lène BRICHLER (Hôpital Avicenne, Bobigny), Duc NGUYEN, Pantxika BELLECAVE, Camille CICCONE, Marie-Edith LAFON (CHU Pellegrin, Bordeaux), Ségolène GREFFE (CHU Ambroise Paré, Boulogne Bill-ancourt), Camille BOUISSE, Nicholas SEDILLOT, Damien BOUHOUR (CH Fleyriat, Bourg en Bresse), Camille CHASSIN (CH Pierre Oudot, Bourgoin-Jallieu), Erwan L’HER, Séverine ANSART, Cécile TRO-MEUR, Dewi GUELLEC (CHRU de Brest), Antoine MERCKX (CH de Cahors), Felix DJOSSOU (CH Andrée Rosemon, Cayenne), Vincent PEIGNE, Carola PIEROBON, Marie-Christine CARRET, Florence JEGO, Margaux ISNARD (CHMS Chambéry NH), Johann AUCHABIE, Roxane COURTOIS (CH de Cholet), Olivier LESENS (CHU Gabriel Montpied, Clermont-Ferrand), Martin MARTINOT (Hôpital Pasteur, Colmar), Olivier PICONE (Hôpital Louis Mourier, Colombes), Marie LACOSTE (CH Alpes Leman Contamine sur Arve), Brigitte ELHAR-RAR, Valérie GARRAIT, Isabelle DELACROIX, Thomas MAITRE, Jean Baptiste ASSIE (Centre Hospitalier Intercommunal de Créteil), Elsa NYAMANKOLLY (CH de Dax), François Xavier CATHERINE, Ma-thieu BLOT, Sophie MAHY, Marielle BUISSON, Lionel PIROTH, Florence GUILLOTIN, Alexis DE ROUGEMONT (CHU de Dijon Bourgogne), Valentine CAMPANA, Jérémie PASQUIER, André CABIE (CHU de Martinique, Fort de France), Simon BESSIS (Hôpital Raymond Poincaré, Garches), Olivier EPAULARD, Nicolas TERZI, Jean-François PAYEN, Laurence BOUILLET, Rebecca HAMIDFAR, Marion Le MARECHAL (CHU de Grenoble), Moïse MACHADO, Audrey BARRELET, Alexandra BEDOSSA (Grand Hôpital de l’Est Francilien site Marne-la-Vallée, Jossigny), Gwenhaël COLIN, Romain DECOURS, Thomas GUIMARD (CHD Les Oudairies, La Roche Sur Yon), Cécile GOUJARD, Stéphane JAUREGUIBERRY, Antoine CHERET (Hôpital Universitaire Bicêtre, Le Kremlin-Bicêtre), Anne Sophie RESSEGUIER (CH Emile Roux, Le Puy en Velay), Julien POISSY, Daniel MATHIEU, Saad NSEIR, Ilka ENGELMANN, Martine REMY (CHRU de Lille, Hôpital Salengro), Fanny VUOTTO, Benoit GACHET, Karine FAURE, Marielle BOYER-BESSEYRE, Kazali Enagnon (CHRU de Lille, Hôpital Fourrier), Marc LAMBERT, Arnaud SCHERPEREEL, Dominique DEPLANQUE, Stéphanie FRY, Cécile YELNIK (CHRU de Lille, Hôpital Albert Calmette), Laurent BITKER, Mehdi MEZIDI, Hodane YONIS, Nicolas BENECH, Thomas PER-POINT, Anne CONRAD, Vinca ICARD, Martine VALETTE (Hôpital de la Croix Rousse, Lyon), Simon-Djamel THIBERVILLE (Hôpital Louis Raffalli, Manosque), Stanislas REBAUDET (Hôpital Européen de Marseille), Bertrand DUSSOL (Hôpital Conception, Marseille), Sylvain DIAMANTIS, Catherine CHAKVEATZE, Clara FLATEAU (Groupe Hospitalier Sud Ile De France, hôpital de Melun), Vincent DINOT, Rostane GACI, Nadia OUAMARA (Hôpital de Mercy, CHR Metz-Thionville, Ars laquenexy), Hajnal-Gabriela ILLES, Louis GER-BAUD MORLAES, Jérôme DIMET (CH de Mont Marsan), Vincent LE MOING, Nathalie PANSU, Clément LE BIHAN, Brigitte MONTES (CHU de Montpellier), Anne Sophie BOUREAU, Clotilde ALLAVENA, Sabelline BOUCHEZ, Romain GUERY, Paul LE TURNIER, Cécile MEAR-PASSARD, Virginie FERRE (CHU de Nantes), Christophe RAPP (Hôpital Américain de Paris, Neuilly sur Seine), Elisa DE-MONCHY, Céline MICHELANGELLI, Karine RISSO (CHU de Nice), Paul LOUBET, Jean-Phillippe LAVIGNE (CHU Caremeau, Nîmes), Etienne DE MONTMOLLIN, Juliette PATRIER, Paul Henri WICKY, Lucie LE FEVRE, Pierre JACQUET, Raphael BORIE, Antoine DOS-SIER, Dominique LUTON, Valentina ISERNIA, Sylvie LE GAC (Hôpital Bichat, Paris), Cécile AZOULAY, Nicolas CARLIER, Liem LUONG, Marie LACHATRE, Odile LAUNAY (Hôpital Cochin, Paris), Younes KERROUMI, Vanina MEYSSONNIER (Groupe Hospitalier Diacon-esses Croix Saint-Simon, Paris), Jean-Luc DIEHL, Jean-Sébastien HULOT, Bernard CHOLLEY, Jean-Benoît ARLET, Olivier SANCHEZ (Hôpital Européen Georges Pompidou, Paris), Victoria MANDA, Laurène AZEMAR, Guylaine CASTOR-ALEXANDRE (Hôpital Lariboisière, Paris), Karine LACOMBE, Thibault CHIARABINI, Bénédicte LEFEBVRE, Laurence MORAND-JOUBERT, Djeneba FOFANA, Aur-élie SCHNURIGER (Hôpital Saint Antoine, Paris), Nathalie DE CAS-TRO, Constance DELAUGERRE, Marie-Laure CHAIX (Hôpital Saint Louis, Paris), Julie CHAS (Hôpital Tenon, Paris), Valérie GABORIEAU, Eve LE COUSTUMIER, Walter PICARD (CH de Pau), Jean-Benoît ZABBE, Florent PEELMAN, Edouard SOUM (CH de Périgueux), Hu-gues AUMAÎTRE (CH de Perpignan), Elodie CURLIER, Rachida OUISSA, Isabelle FABRE (CHU de Pointe à Pitre, Guadeloupe), Blandine RAMMAERT (CHU de Poitiers), Nadia SAIDANI (CH Quimper), Firouzé BANI-SADR, Maxime HENTZIEN, Yohan N’GUYEN, Juliette ROMARU, Kévin DIDIER (CHU Reims), Isabelle ENDERLE, Vincent THIBAULT, Fabrice LAINE, Matthieu LESOU-HAITIER, Matthieu REVEST, Pierre TATTEVIN (CHU Rennes), Jean-Christophe PLANTIER, Manuel ETIENNE, Véronique LEMEE, Eglan-tine FERRAND DEVOUGE, Kévin ALEXANDRE, Elise ARTAUD-MACCARI (CHU Rouen), Nathalie ALLOU, Marie LAGRANGE, Julien JABOT (CH Saint Denis et Saint Pierre, La réunion), Benoît ROZE, Delphine BREGEAUD, Younes AIT TAMLIHAT (CH Saintes), Ali HACHEMI (CH Soisson), Hélène SALVATOR, Erwan FOURN, David ZUCMAN (Hôpital Foch, Suresnes), Eric DELAVEUVE, Coline JAUD-FISCHER, Paul DUNAND (Hôpital Bel Air, Thionville), François BIS-SUEL (CH Thonon les Bains), Karen DELAVIGNE, Marie PIEL-JULIAN, Pierre DELOBEL, Benjamine SARTON, Marie RAFIQ, Guil-laume MARTIN-BLONDEL, Laurent GUILLEMINAULT, Marlène MURRIS, Agnès SOMMET (CHU de Toulouse), Eric SENNEVILLE, Olivier ROBINEAU, Agnès MEYBECK (CH Tourcoing), Denis GAROT, Laurent PLANTIER, Valérie GISSOT, Julien MARLET, Karl STEFIC, Catherine GAUDY-GRAFFIN, Francis BARIN, Adrien LEMAIGNEN (CHU Bretonneau, Tours), Grégory CORVAISIER, Delphine LARIVIERE, Marie LANGELOT-RICHARD (CH Vannes), Elisabeth BOTELHO-NEVERS, Amandine GAGNEUX-BRUNON, Tiffany TROUILLON, Thomas BOURLET, Thomas BOURLET, Sylvie PILLET, Bruno POZZETTO (CHU de St Etienne), Antoine KIMMOUM, Bruno LEVY, Mathieu MATTEI, François GOEHRINGER, Christian RABAUD, Sibylle BEVILACQUA, Benjamin LEFEVRE, Anne GUIL-LAUMOT, Véronique VENARD, Hélène JEULIN, Evelyne SCHVOERER, Cédric HARTARD (CHU de Nancy, Vandoeuvre les Nancy), Pauline CARAUX PAZ, Laurent RICHIER, Mohamed EL SANHARAWI (CH Villeneuve St Georges).

